# Metabolites mediate genetic effects on disease in the Canadian Longitudinal Study on Aging

**DOI:** 10.1101/2025.11.12.25339921

**Authors:** Joosung Min, Olga Vishnyakova, Angela Brooks-Wilson, Lloyd T. Elliott

**Affiliations:** Department of Statistics and Actuarial Science, Simon Fraser University, 8888 University Dr., Burnaby, V5A 1S6, British Columbia, Canada; Department of Biomedical Physiology and Kinesiology, Simon Fraser University, 8888 University Dr., Burnaby, V5A 1S6, British Columbia, Canada; Department of Basic and Translational Research, BC Cancer, 675 West 10th Ave, Vancouver, V5Z 0B4, British Columbia, Canada

**Keywords:** causal mediation analysis, genetic association, aging, metabolomics, Canadian Longitudinal Study on Aging, CLSA

## Abstract

Understanding the biological mechanisms linking genetic variants to disease risk is essential for advancing precision health. We have developed a causal mediation analysis framework, the C-MAPLE (Causal Mediation Analysis of Pathways Linking Exposures) method, to identify disease-causing pathways for which the effect of genetic variants is mediated through metabolites to impact age-related diseases. Unlike Mendelian randomization, our approach is robust to horizontal pleiotropy, and models multiple mediators and interactions between genetic variants and metabolites simultaneously. To ensure robust model selection, we incorporate least absolute shrinkage and selection operator (LASSO) with stability selection, which can effectively select relevant mediators even in the presence of unmeasured confounding. We also introduce a dynamic adjustment to the number of bootstrap trials to reduce computational burden during uncertainty estimation. Applying this novel framework to the Canadian Longitudinal Study on Aging, we identified 190 potential causal links involving 108 genetic variants, 176 metabolites, and 6 age-related diseases. Our method and findings highlight the utility of causal mediation analysis in uncovering metabolite-mediated genetic mechanisms. This method, combined with large-scale population data sets, has the potential to revolutionize the identification of targets for downstream clinical research, and the development of personalized disease prevention, interventions, and therapeutics.

## 1 Introduction

Genome-wide association studies (GWAS) have been used extensively to understand disease genetics, leading to the discovery of many disease-causing genetic variants [1]. These genetic variants and the genes they encode have provided insights into risk prediction and disease mechanisms [2–5], thereby enhancing clinical care [6–9]. In addition, recent technological advancements, such as mass spectrometry and nuclear magnetic resonance (NMR) spectroscopy, have enabled researchers to measure the metabolome. The metabolome refers to the set of small molecules, known as metabolites, found in a biological sample. These metabolites are the products of cellular processes, and include amino acids, sugars, lipids, and nucleotides. Studying the metabolome helps us understand the biochemical activities and physiological state of an organism, providing insights into health, disease, and the effects of exposures [10]. GWAS conducted on metabolite phenotypes have contributed significantly to understanding the genetic basis of metabolite variation, which in turn can provide insights into biological processes related to disease etiology [11,12].

Recently, work has been done to extend GWAS protocols to test for causal relationships. One such notable method is Mendelian randomization (MR). MR utilizes genetic variants as *instruments* (a variable that may cause a change in the outcome through the exposure variable) to determine whether an observational association between a risk factor and an outcome is consistent with a causal effect [13]. This method takes advantage of the natural random assortment of genetic variants from parents to offspring. Through MR, researchers have identified potential causal connections across various fields such as behavioural science [14], socioeconomics [15,16], environmental science [17], and biomedical science [18–21], including links between metabolites and diseases [22].

Mediation analysis aims to uncover the mechanisms behind an observed relationship between an exposure and an outcome by introducing a third variable, known as a mediator variable [23]. In mediation analysis, the causal effect of an exposure on an outcome, inclusive of any effects mediated through potential mediators, is referred to as the *total effect*. This effect can be partitioned into two components. One component is the *direct* effect of the exposure on the outcome, which bypasses the mediators, and the other component is the *indirect* effect, which is the effect of the exposure operating through the mediators [24]. MR methodologies have been employed to estimate indirect effects, and have been extended to model multiple mediators simultaneously, including two-step MR [25] and network MR [26]. Examples of Mendelian randomization (MR) applications include assessing how glycaemic and lipid risk factors mediate the effect of body mass index (BMI) on coronary heart disease (CHD), utilizing genetic variants linked to BMI as instruments [27]. However, the estimation of mediation effects using MR-based approaches is limited by several factors [28]. One key assumption in MR is that the genetic variant, typically represented by a single-nucleotide polymorphism (SNP), affects the outcome solely through the mediator, without exhibiting horizontal pleiotropy, a condition often referred to as the *exclusion restriction* [29]. Also, most MR approaches assume no interactions between the exposure and the mediator. Furthermore, extensions to multiple mediators are methodologically complex and require independent genetic instruments for each mediator, which are unavailable when the genetic variants are correlated [24,30].

An alternative framework for estimating indirect effects is provided by causal mediation analysis [31,32]. Unlike MR, causal mediation analysis methods do not rely on strong instrumental variables or the assumption of no horizontal pleiotropy. Additional advantages of causal mediation analysis include greater flexibility in modeling multiple mediators using regression-based approaches and the ability to explicitly adjust for measured covariates, including potential interactions among metabolites, or between metabolites and genetic variants [32–36]. Because of these advantages, causal mediation analysis has been adopted in numerous recent studies [37–42]. However, causal mediation analysis also relies on strong assumptions: 1) that there are no unmeasured confounders, and 2) that the models for exposure, mediator, and outcome are all correctly specified. While both assumptions are hard to address, the first assumption can be validated through sensitivity analysis [43].

In this study, we developed C-MAPLE (Causal Mediation Analysis of Pathways Linking Exposures) framework. C-MAPLE integrates a weighting-based causal mediation approach [33] with stability variable selection with LASSO [44,45]. The weighting-based approach mitigates bias that may arise from misspecification of mediator models, while stability selection procedure enables reliable identification of key mediators even in the presence of unmeasured confounding. Our simulation studies demonstrated that this combination yields robust model specification.

We applied this framework to data from the Comprehensive cohort of the Canadian Longitudinal Study on Aging (CLSA; [46]), uncovering 190 potentially causal associations among genetic variants, metabolites, and age-related diseases, based on estimates of total indirect effects. Many of these triplets (comprised of a genetic variant, a metabolite, and a disease) involved multiple metabolites associated with a SNP. For each triplet, we report the total indirect effect along with uncertainty estimates obtained via bootstrapping, where the number of iterations was dynamically adjusted to balance computational efficiency and statistical stability. Our novel approach offers a flexible and robust framework for identifying causal pathways through indirect effect estimation, while addressing potential sources of bias and reducing computational burden.

The remainder of this manuscript is organized as follows. Section 2 presents the results of the analyses, including both the simulation study and the application of our framework to the CLSA data. Section 3 summarizes the findings and discusses their implications. Section 4 provides background on causal mediation analysis, outlining key notation, the conventional regression-based approach, the weighting-based method that serves as the basis for our analysis, and the dynamic bootstrapping procedure. Section 5 and 6 contain supplementary information, including acknowledgements, author contributions, funding information, and data availability statements.

## 2 Results

In this section, we present the results from simulation studies and the application of the C-MAPLE framework to the CLSA cohort. We used simulation to evaluate the ability of stability selection with LASSO to identify relevant mediators in the presence of irrelevant ones, under varying levels of unmeasured confounding and different sample sizes. Findings indicate that, with a sufficiently high selection threshold, the method can reliably identify true mediators even when the outcome is strongly influenced by unmeasured confounders. The simulation study result is illustrated in Figure 2.

**Figure 1.**
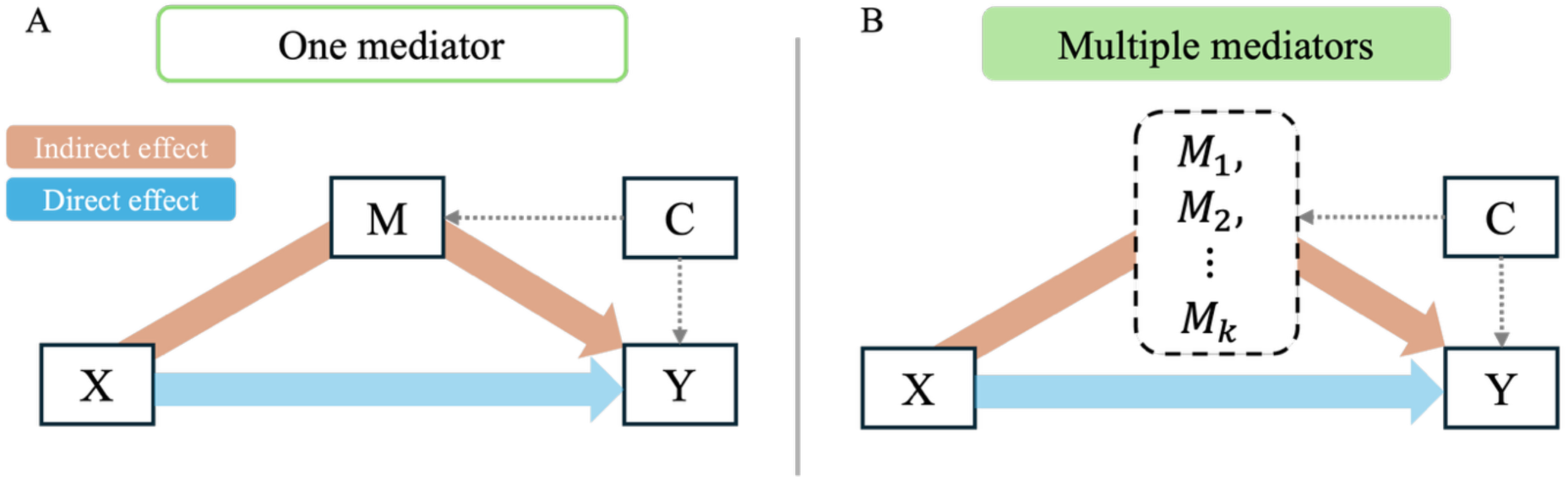
Illustration of the causal relationship between exposure **X**, mediators **M**, and outcome **Y** variables, in the presence of confounders (C), shown for the case of a single mediator (A) and multiple mediators (B).

**Figure 2.**
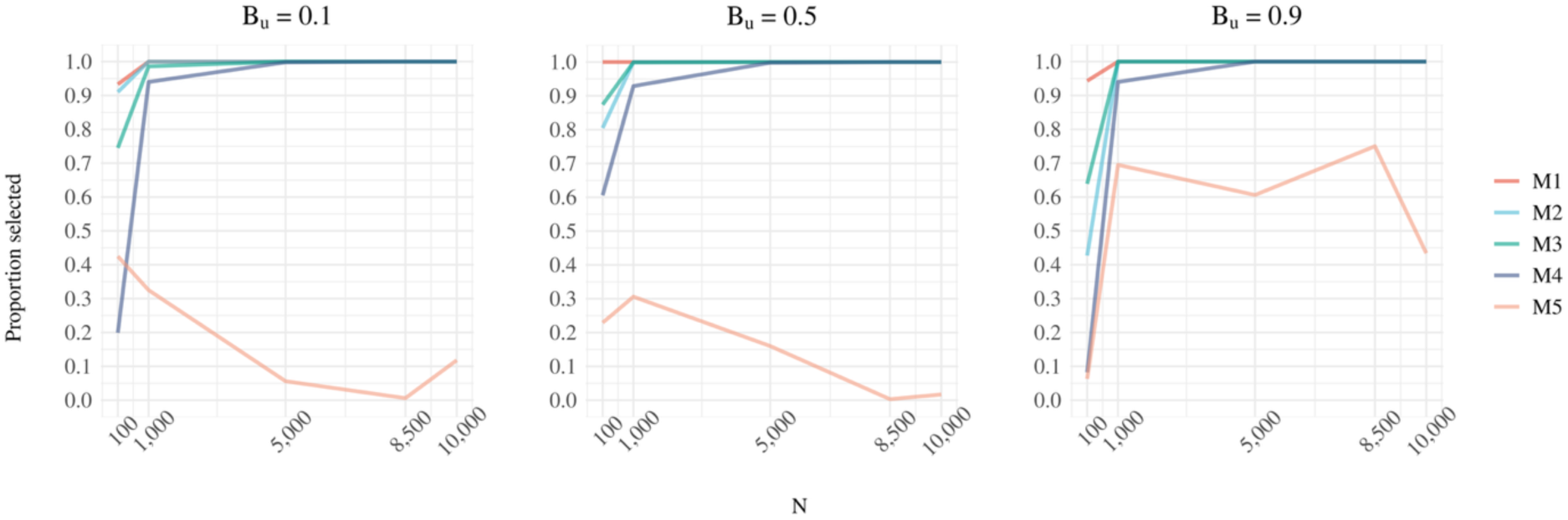
Proportions of individual mediators selected across 1,000 trials using stability variable selection with LASSO are shown under varying effect sizes of the unobserved confounder U. Among the mediators, only M_1_, M_2_, M_3_, and M_4_ are truly causally associated with the outcome Y, while M_5_ is associated with the exposure X, but not causally linked to Y. The values β_u_ = 0.5 and 0.9, represent scenarios where the effect of U is equal to or greater than those of M_2_, M_3_ and M_4_. With a high selection threshold (≥ 0.8), stability selection with LASSO consistently selected the true mediators (M_1_ to M_4_) more frequently than the non-relevant mediator M_5_.

When applied to the Comprehensive Cohort of the Canadian Longitudinal Study on Aging (CLSA-COM; [46]), the C-MAPLE method identified 190 potential causal associations involving 108 genetic variants, 176 metabolites, and 6 age-related diseases. Per disease, The mediating metabolites identified are classified by super-pathways as follows: 97 Lipid super-pathway, 52 Amino Acid, 2 Carbohydrate, 5 Cofactors and Vitamins, 1 Energy, 4 Nucleotide, 4 Partially Characterized Molecule, 2 Peptide, and 9 Xenobiotics. Our method and findings highlight the utility of causal mediation analysis in uncovering metabolite-mediated genetic mechanisms. The full list of identified triplets and their NIEs are available in Supplementary Table 1.

### 2.1 Simulation study for stability variable selection with LASSO

We conducted a simulation study to evaluate the performance of LASSO regression with stability selection in identifying true causal mediators. Specifically, we assessed its ability to select relevant mediators (*M*_1_ to *M*_4_) in the presence of an irrelevant mediator (*M*_5_), across varying levels of unmeasured confounding (*β_u_* ∈ 0.1,0.5,0.9) and different sample sizes (*N* ∈ 100,1000,5000,8500,10000). The *β_u_* values of 0.5 and 0.9 represent scenarios in which the effect of the unmeasured confounder equals or exceeds the effects of some relevant mediators (*M*_2_ to *M*_4_). Further details of the data generation process are provided in the Methods section. Figure 2 displays the selection frequency of each mediator across 1,000 bootstrap iterations.

The results show that, with a sufficiently high selection threshold (≥ 0.8), stability selection with LASSO consistently identifies truly relevant mediators, even in the presence of substantial unmeasured confounding.

### 2.2 Causal mediation analysis on the CLSA cohort

We applied the C-MAPLE framework to the Comprehensive Cohort of the Canadian Longitudinal Study on Aging (CLSA-COM). Figure 3 shows the estimated natural indirect effects (NIEs) on the risk ratio scale, expressed as base-10 logarithms (log10(NIE)), together with their 95% bootstrap confidence intervals. Positive log10(NIE) values indicate that the presence of the effect allele (A1) increases disease risk, whereas negative values indicate a reduction in risk relative to the reference allele (REF) at the SNP. Figure 3A displays the log10(NIE) values by SNP. A single SNP may exhibit indirect effects on multiple diseases. For example, rs10774021 on chromosome 12 showed statistically significant log10(NIE) values for Parkinson’s disease, Osteoporosis, and Type-2 diabetes (T2D). In addition, multiple NIE estimates may exist for a single SNP–disease pair because each was evaluated under three distinct genotype comparisons: homozygous reference vs. heterozygous (*test01*), homozygous reference vs. heterozygous effect (*test02*), and heterozygous vs. homozygous effect (*test12*). Figure 3B and 3C summarize the results by mapped genes and mediating metabolites, respectively.

**Figure 3.**
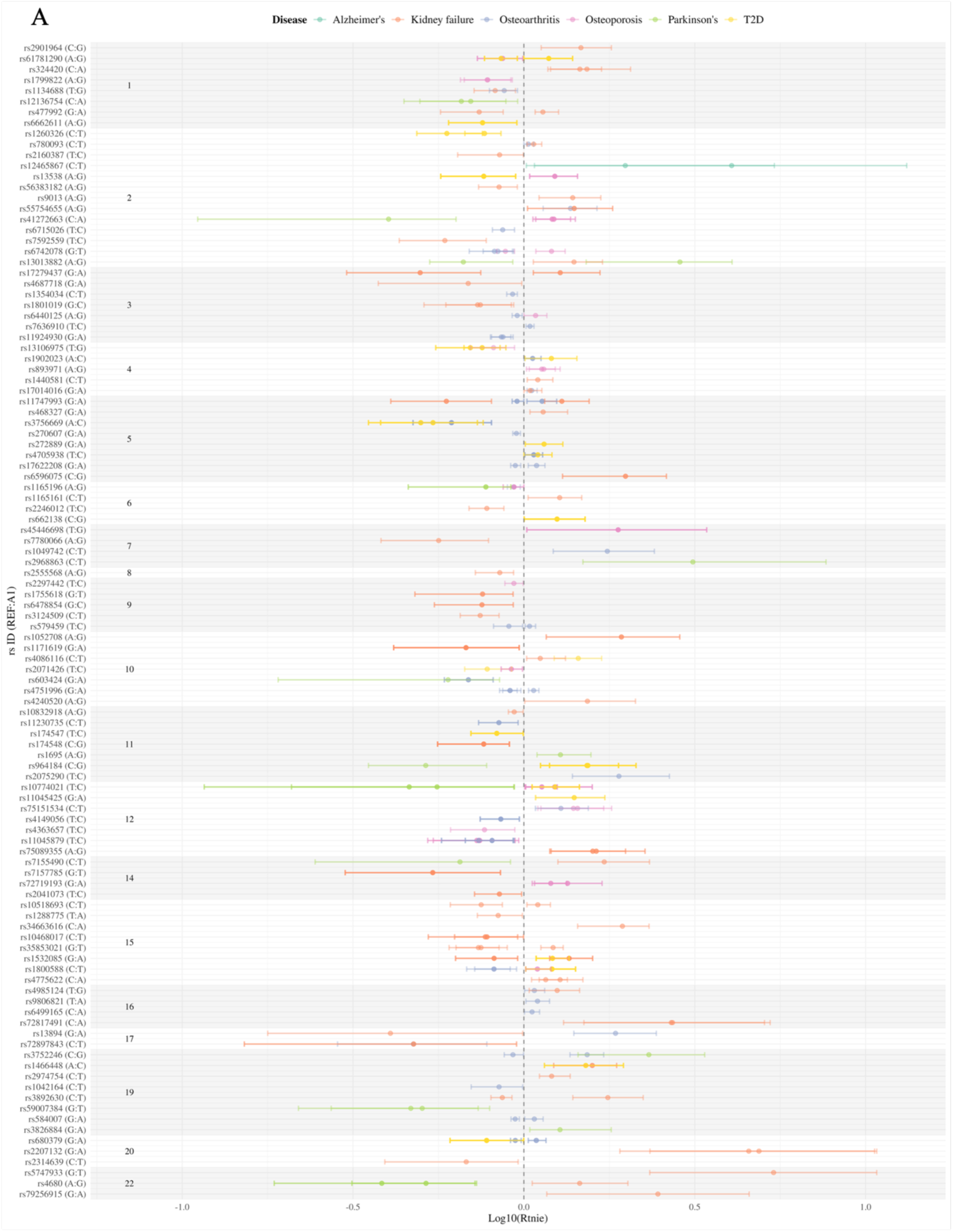
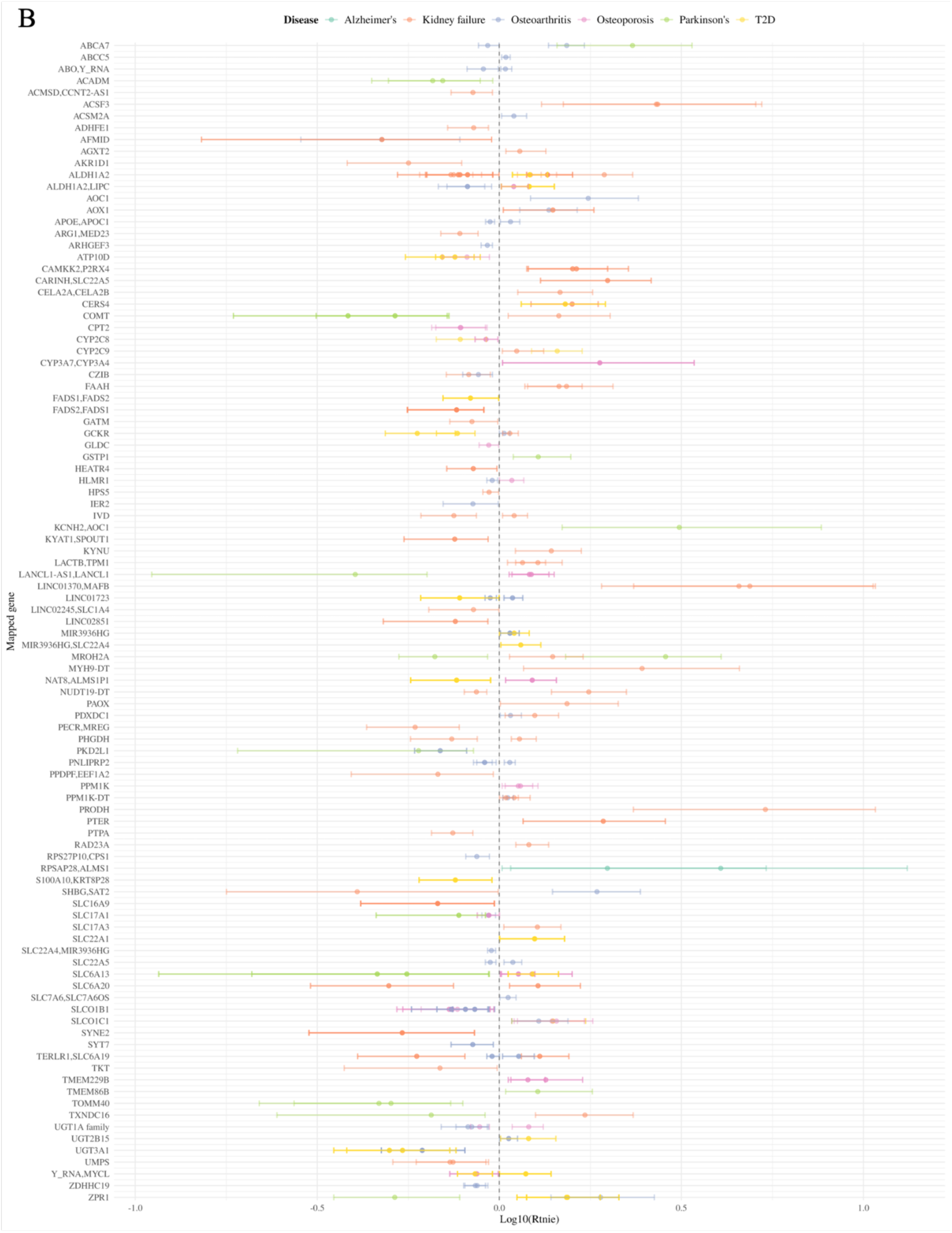
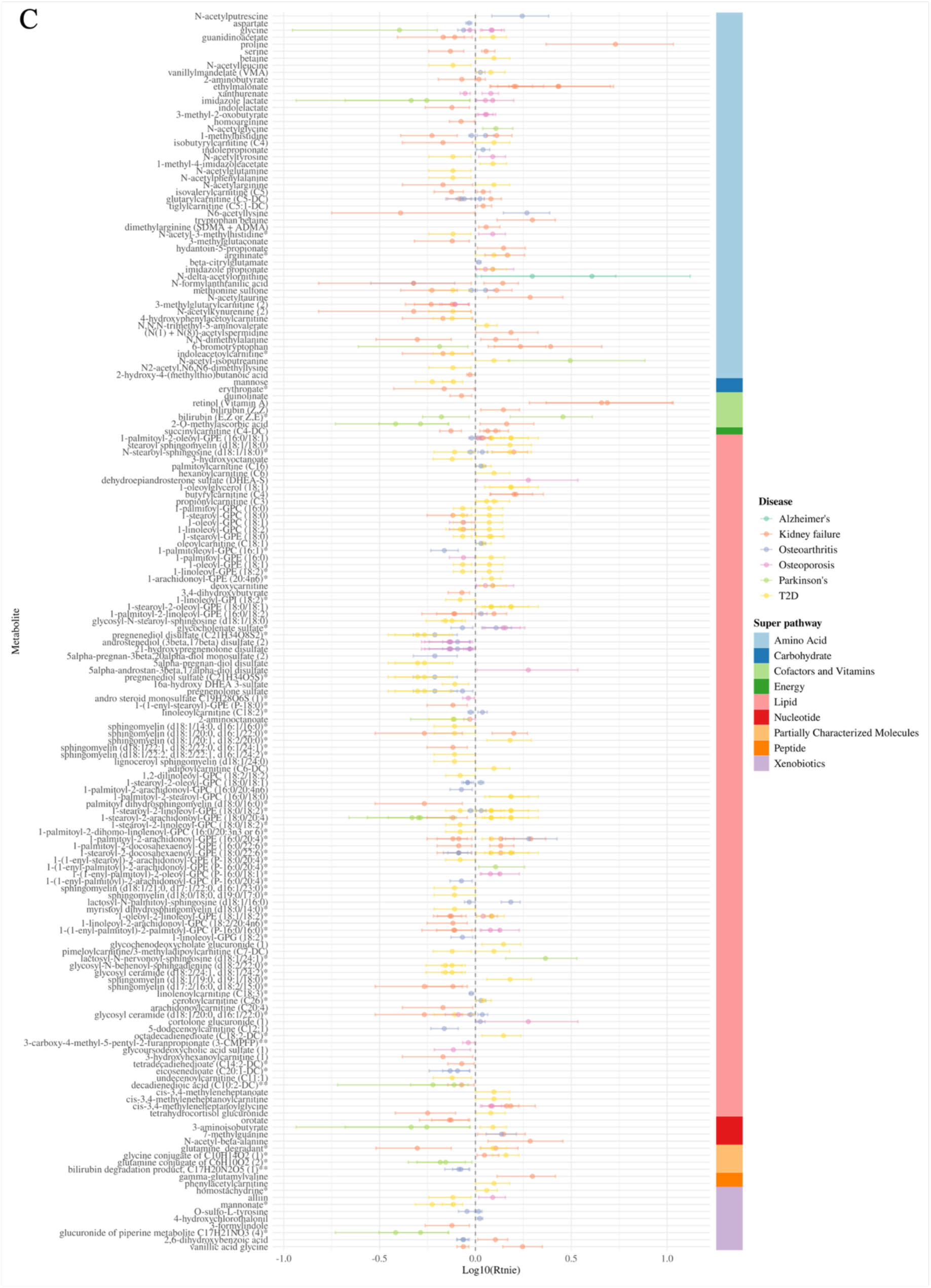
Estimated natural indirect effects (NIEs) on the risk ratio scale, expressed as base-10 logarithms (log10(NIE)), with 95% bootstrap confidence intervals. Positive values indicate that the effect allele (A1) increases disease risk, and negative values indicate reduced risk relative to the reference allele (REF). **A.** log10(NIE) values by SNP, where a single SNP may affect multiple diseases (e.g., rs10774021 for Parkinson’s disease, osteoporosis, and T2D) or show multiple estimates for the same disease from three genotype comparisons. **B.** Results summarized by mapped genes. **C.** Results summarized by mediating metabolites.

We report NIE, NDE, and E-values for individual triplets in Supplementary Table 1. For example, the rs6715026 SNP on chromosome 2 (reference allele T, risk allele C) showed an NIE of 0.867 when comparing heterozygous to homozygous minor genotypes *(test12)*, suggesting that carrying two copies of the risk allele, rather than one, may be advantageous. In our GWAS of metabolites (Supplementary Table 2), this SNP was positively associated with glycine levels (β = 0.0135, adjusted *p* = 4.51 × 10⁻⁶), indicating that an increase in glycine induced by the risk allele may reduce the risk of osteoarthritis. Furthermore, a statistically significant NDE for this triplet suggests the presence of an additional pathway, independent of glycine, through which the SNP may influence osteoarthritis risk.

Below, we highlight examples of variant–metabolite–disease triplets that may reflect underlying causal pathways. We selected these triplets for this discussion based on two criteria: 1) the presence of multiple mediators, and 2) the existence of previous literature noting the biological relevance of the variant or metabolite to the disease. While our data-driven approach does not establish definitive causation, their consistency with known biology supports the effectiveness of this approach for identifying and understanding causal pathways. New associations identified by this method offer valuable hypotheses that can guide and refine the focus of future experimental or mechanistic studies.

#### 2.2.1 Association of rs10774021 *(SLC6A13)* with multiple diseases

Our analysis suggests that the effect of the rs10774021 polymorphism in *SLC6A13* on multiple diseases (kidney failure, osteoporosis, and type 2 diabetes [T2D]), may be mediated by the metabolites guanidinoacetate, 3-aminoisobutyrate, deoxycarnitine, imidazole propionate, and imidazole lactate. *SLC6A13* encodes a transporter for guanidinoacetate, a precursor of creatine, which is essential for energy metabolism [47]. Because creatine biosynthesis primarily occurs in the kidneys and liver, altered *SLC6A13* function may affect metabolite levels, with downstream effects on muscle function, glucose regulation, and bone health. Previous studies support this mechanistic link. For example, high plasma creatine levels have been associated with increased risk of T2D in men [48]. Mice lacking the ability to produce creatine or guanidinoacetate are protected from diet-induced metabolic syndrome [49]. Imidazole propionate (ImP) has been implicated in both diabetes and cardiovascular disease [50]. Furthermore, ImP disrupts bone homeostasis by inhibiting AMP-activated protein kinase (AMPK) signaling and activating p38*γ* phosphorylation [51]. Genome-wide association studies (GWAS) have shown that the SNP rs10774021 is associated with blood levels of guanidinoacetate [52], 3-aminoisobutyrate [53], deoxycarnitine [22], and various imidazole compounds including 1-methylimidazoleacetate, 1-methyl-4-imidazoleacetate, imidazole lactate, and 1-ribosyl-imidazoleacetate (X-11334) [52,53], sodium bicarbonate levels, body mass index (BMI) [54], waist circumference [55], estimated glomerular filtration rate, and chronic kidney disease [56]. Our findings align with these previous GWAS results and further support a potential metabolic pathway linking rs10774021 to multiple disease outcomes through these metabolites.

#### 2.2.1 Association of rs2968863 *(KCNH2/AOC1)* with Parkinson’s disease

Our results indicate that the rs2968863 SNP, located between the *KCNH2* and *AOC1* genes, may influence the risk of Parkinson’s disease through its effect on N-acetylisoputreanine levels. While *KCNH2* primarily encodes a potassium channel [57], and rs2968863 has been associated with QT interval duration in GWAS studies [58], the SNP’s proximity to *AOC1* which encodes diamine oxidase, an enzyme responsible for putrescine degradation [59,60], suggests a potential regulatory role. This may suggest a potential regulatory effect of this SNP on both genes, or linkage between rs2968863 and SNPs that affect the function of *AOC1*. GWAS findings have associated variants in *AOC1* with levels of N-acetyl-isoputreanine [52], a metabolite in the polyamine pathway [61]. It is therefore plausible that rs2968863 modulates

*AOC1* expression or activity, thereby altering putrescine metabolism and influencing N-acetyl-isoputreanine levels. Dysregulation of polyamine metabolism has been implicated in neurodegenerative diseases such as Parkinson’s disease [62], and N-acetyl-isoputreanine has also been associated with cognitive function in other neurodegenerative contexts [63]. These findings support a potential mechanistic link between rs2968863, polyamine metabolism, and Parkinson’s disease risk.

#### 2.2.2 Association of rs4680 (*COMT*) with Parkinson’s disease and kidney failure

Our findings indicate that the rs4680 polymorphism in the *COMT* gene exerts a significant mediation effect through 2-O-methylascorbic acid (2-O-MA) in the pathogenesis of Parkinson’s disease. *COMT* is involved in catecholamine metabolism, which plays a crucial role in neurotransmitter regulation and may influence neurodegenerative diseases such as Parkinson’s disease [64]. 2-O-MA is a derivative of vitamin C (ascorbic acid), produced via methylation of L-ascorbic acid by *COMT* [65]. While ascorbic acid is well-studied for its essential roles in human health, much less is known about the specific functions or physiological effects of its methylated derivatives like 2-OMA. Although 2-O-MA is structurally similar to vitamin C, methylation may reduce its effectiveness as an antioxidant, since the hydroxyl groups are crucial for ascorbic acid’s redox activity [66,67]. Altered *COMT* activity due to rs4680 may modify 2-O-methylascorbic acid levels, potentially affecting antioxidant capacity. Given the vulnerability of dopaminergic neurons to oxidative damage [68], this mechanism provides a biologically plausible link between rs4680, 2-O-MA, and Parkinson’s disease risk. A similar pathway may also be relevant in renal pathophysiology, where oxidative stress plays a central role [69].

#### 2.2.3 Association of rs1260326 *(GCKR)* with type 2 diabetes

The rs1260326 variant in *GCKR* is associated with T2D risk through its effect on mannose and mannonate metabolism. *GCKR* encodes the glucokinase regulatory protein, which modulates glucokinase activity in response to glucose levels [70]. Several studies have associated this SNP with altered levels of mannose and mannonate [71,72]. Mannose has been associated with insulin resistance and increased diabetes risk [73], while mannose and mannonate were identified as part of a 10-metabolite predictive metabolic signature for type 2 diabetes [74]. The same variant has also been associated with dyslipidemia, including elevated triglycerides and cholesterol [75], which are frequently observed in individuals with metabolic syndrome and diabetes [76]. These observations together suggest that rs1260326 contributes to T2D pathogenesis through mannose and mannonate metabolism and other metabolic pathways.

#### 2.2.4 Other notable results

Most of the indirect effects identified showed consistent directional patterns across comparisons involving different numbers of minor alleles. That is, all natural indirect effects (NIEs) for *test01* (homozygous major vs. heterozygous), *test02* (homozygous major vs. homozygous minor), and *test12* (heterozygous vs. homozygous minor) were either all greater than 1 or all less than 1.

However, some variant–metabolite–disease triplets exhibited potential non-additive effects of minor alleles. One example is the association between rs477992 and kidney failure, mediated by serine. The rs477992 SNP is located in intron 1 of the PHGDH gene, which encodes phosphoglycerate dehydrogenase, a key enzyme in the serine biosynthesis pathway [77]. rs477992 is associated with plasma serine levels in GWAS studies [78]. Our results indicate that the NIE comparing homozygous major alleles to heterozygous alleles was 1.136 (95% CI: 1.079, 1.263), whereas the NIE comparing heterozygous alleles to homozygous minor alleles was 0.74 (95% CI: 0.57, 0.87). These findings suggest that carrying a single copy of the minor allele may have a more deleterious effect on kidney function via serine than carrying two copies, indicating a possible heterozygote-specific effect at this locus.

Other triplets with potential heterozygote-specific effects include those involving the following SNPs: rs17279437, rs11747993, rs35853021, rs1532085, and rs3892630 (kidney failure); rs17622208, rs579459, rs4751996, rs3752246, rs584007, and rs680379 (osteoarthritis); rs6742078 (osteoporosis); rs13013882 (Parkinson’s disease); and rs61781290 and rs174547 (type-2 diabetes).

As our study is purely data-driven, these results should be interpreted with caution. Further biological studies or complementary approaches will be necessary to confirm and extend these observations.

## 3 Discussion

We investigated the potential of a causal mediation analysis-based framework for identifying causal mechanisms linking genetic variants, metabolites, and disease risks. CMA distinguishes between the overall effect of an exposure and its direct and indirect components, where the indirect effect captures the impact of the exposure on the outcome as mediated through intermediate variables. Unlike Mendelian randomization, causal mediation analysis is not biased or invalidated by horizontal pleiotropy. The weighting-based mediation analysis we apply in this work uses an inverse probability weighting approach that reduces the risk of biased estimates. This method requires only two models (exposure and outcome) to be correctly specified, in contrast to regression-based approaches, which demand correct specification of models for all mediators.

To improve model selection, we employed stability variable selection with LASSO, enhancing robustness. Our simulation study shows that stability selection with LASSO can effectively exclude irrelevant mediators, even in the presence of unmeasured confounders whose effects exceed those of some relevant mediators.

We applied this approach to the CLSA Comprehensive dataset and conducted tests to identify significant indirect effects using a non-parametric bootstrap procedure. The number of bootstrap trials was dynamically adjusted to significantly reduce the computational burden. As a result, our analysis revealed 190 potential causal links involving 108 genetic variants and 176 metabolites. Many of these links include multiple metabolites as mediators and are supported by their known biological functions.

Our approach has several limitations. Weighting-based mediation analysis estimates the joint indirect effect of all mediators rather than assessing each individually. Evaluating mediators one at a time would lead to biased estimates due to outcome model misspecification. Additionally, all models in this study assume linear relationships between variables. However, the flexibility of weighting-based causal mediation analysis allows for the incorporation of non-linear models.

Our framework offers a valuable toolset for uncovering data-driven evidence of causal mechanisms between genetic variants and age-related diseases, with metabolites serving as mediators. Combined with large population-based metabolomics data sets, the C-MAPLE method has the potential to provide substantial insights into the genetic and molecular basis of health and disease. Building on our previous work that identified “Sweet Spots” in phenotypic and metabolomic biomarkers, which represent optimal ranges associated with healthy aging, the present framework offers a natural extension. Our future work will integrate Sweet Spot biomarkers into causal mediation analyses to estimate pathways and quantify the indirect effects of deviations from these optimal ranges on aging-related outcomes. Together, data-driven Sweet Spot identification and robust mediation analysis form a unified strategy for elucidating mechanisms that promote healthy aging.

## 4 Methods

In this section, we describe the data and methodologies used in this study. This includes a brief overview of the Comprehensive Cohort of the Canadian Longitudinal Study on Aging (CLSA), an introduction to causal mediation analysis, and a description of the stability selection method with LASSO. We also outline the dynamic bootstrap procedure, the synthetic data generation process used in the simulation study, and the quality control steps applied to the CLSA-COM dataset.

### 4.1 Canadian Longitudinal Study on Aging Comprehensive Cohort

Our selected subset for analysis from the Canadian Longitudinal Study on Aging Comprehensive Cohort (CLSA-COM; [46]) comprised 30,097 participants aged between 45 and 85 years. Of these participants, 52.3% were female, and the average age was 63.1 years at time of the blood draw for metabolomic measurements. 94% of participants self-reported as belonging to the White ethnic group, followed by Asian (2%), and Black (0.7%). Their genotypes (single-nucleotide polymorphisms; SNPs), metabolites, and lifestyle factors (alcohol consumption, smoking frequency, and body-mass index) were included. After a series of quality control steps, genome and metabolome data from 8,698 subjects were used for analyses. The details about the quality control steps are described in Section 4.6.

### 4.2 Genome-wide association study on metabolites

In order to conduct causal mediation analysis, an association between the exposure (genetic variations) and mediators (metabolites) must exist. To establish these associations, we performed genome-wide association studies across the 193 metabolites (mGWAS). We identified a total of 1,112 significant associations across 679 SNPs and 183 metabolites that passed a Bonferroni-adjusted *p*-value threshold of 0.05*/*(616,294 ∗ 193) = 4.20 × 10^−10^. mGWAS were conducted using *PLINK 2.0* [79,80], adjusting for covariates including age, sex, the first 10 genetic principal components, genotyping batch, and hours since the last meal or drink. 304 of these SNPs were identified as lead SNPs using the *peaks* algorithm [81,82]. Consequently, 483 target SNP-metabolite-disease triplets were selected for causal effect estimations.

### 4.3 Causal mediation analysis

In this section, we provide background on the concept of causal mediation analysis, including explanations of key notations. We then briefly introduce the conventional regression-based mediation analysis, outlining its strengths and limitations. Finally, we describe the weighting-based mediation approach, which addresses the limitations of the regression-based method and serves as the basis for our analysis.

#### 4.3.1 Potential outcome framework

Let *X* denote an exposure of interest, *M* a set of potential mediators, *Y* an outcome, and *C* a set of measured covariates that are not affected by the exposure. In the case of a single mediator, the relationships between *X*, *M*, *Y*, and *C* are illustrated as in Figure 1A. To define the causal mediation effects, we use the potential outcome framework [32,33,83]. Let *Y_a_* denote a subject’s potential outcome under an exposure status *x* (this exposure status may be contrary to the actual exposure status). Let *M_x_* denote a subject’s counterfactual value of *M* that would be observed if *X* were set to the value *x*. Then, the *natural direct effect* (NDE) and the *natural indirect effect* (NIE) of exposure *X* on the outcome *Y* is defined as:

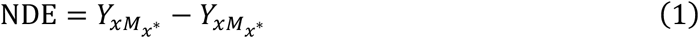

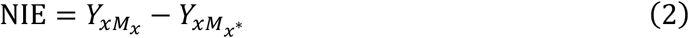

The NDE compares the effect of *X* = *x* versus *X* = *x*^∗^ while setting *M* to what it would have been under exposure status *X* = *x*^∗^. The NIE compares two hypothetical outcomes: one where the mediator *M* is set to the value it would have taken if *X* = *x*, and another where *M* is set to the value it would have taken if *X* = *x*^∗^. The *total effect* (TE) *Y_x_* − *Y_x_*_∗_ can thus be expressed as:

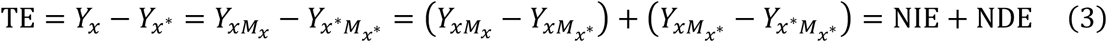

With these equations, we can define the average natural direct and indirect effects conditional on *C* as: E[*Y_xMx_*_∗_ − *Y_x_*_∗_*_Mx_*_∗_|*c*] and E[*Y_xMx_* − *Y_xMx_*_∗_|*c*], respectively [33].

#### 4.3.2 Regression-based mediation analysis

In a conventional regression-based mediation analysis, two types of models are defined: mediator, and outcome models [33]. These models are described as follows:

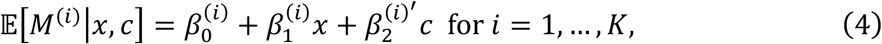

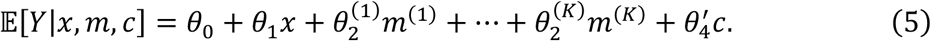

These models assume that there are no exposure-mediator or mediator-mediator interactions. Then, the average NDE and NIE can be defined as:

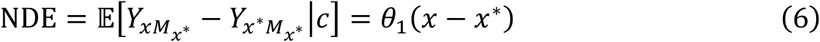

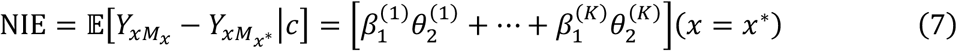

For a binary outcome, the outcome model in Equation 5 can be replaced with the following:

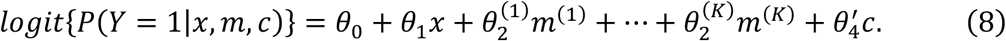

Then, the log of the NDE and NIE odds ratios are given as follows:

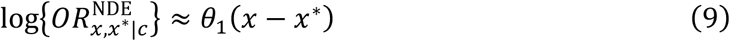

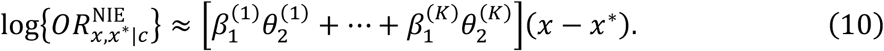

Misspecification of any of these models can lead to biased causal effect estimations [30,33]. As the number of mediators increases, more mediator models need to be correctly specified (and so do potential interaction terms between the variables), adding to the risk of bias. This limitation underscores the need for a method that can flexibly accommodate multiple mediators without requiring correct specification of individual mediator models.

#### 4.3.3 Weighting-based mediation analysis

Weighting-based mediation analysis offers greater flexibility compared to regression-based methods [33]. Similar to the regression approach, the weighting-based approach also assumes that the models are correctly specified, and misspecification of either model will result in biased estimation. However, the weighting-based approach requires only the correct specification of the exposure and outcome models, which can reduce the risk of bias due to model misspecification in scenarios with multiple mediators. In contrast, the regression approach requires correct specification of the outcome model and also all mediator models. The weighting-based approach estimates the marginal NDE and NIE, defined as follows:

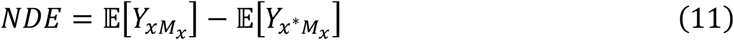

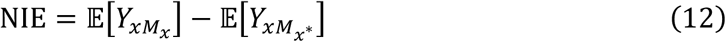

These effects can also be described as risk ratios (RR) as follows:

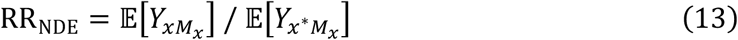

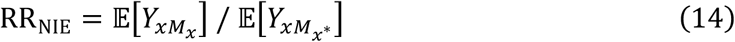

Estimating these causal effects requires computing three counterfactuals: E[*Y_xMx_*], E[*Y_x_*_∗_*_Mx_*_∗_], and E[*Y_x_*_∗_*_Mx_*]. The first two counterfactuals E[*Y_xMx_*] and E[*Y_x_*_∗_*_Mx_*_∗_] are obtained by computing weighted averages of subjects with *X* = *x* and *X* = *x*^∗^, respectively. Each subject *i* is assigned weights *P*(*X* = *x*)*/P*(*X* = *x*|*c_i_*) and *P*(*X* = *x*^∗^)*/P*(*X* = *x*^∗^|*c_i_*). Here *c_i_* represents the actual covariate value for subject *i*. The probabilities *P*(*X* = *x*|*c_i_*) and *P*(*X* = *x*^∗^|*c_i_*) can be computed using an exposure model. We use the following logistic regression model:

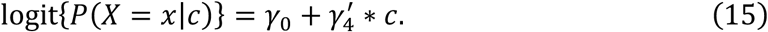

For the counterfactual E[*Y_xMx_*_∗_], an outcome model, as illustrated in Equation 5 or Equation 8, can be fitted depending on the outcome type. The outcome model can include exposure-mediator or mediator-mediator interactions. Then, the potential outcomes for subjects with *X* = *x*^∗^ are obtained while setting their *X* at *x* instead of at *x*^∗^, but using their own values of the mediators *M* = *m_i_*, and covariates *C* = *c_i_*. After obtaining the predicted values, the counterfactual estimate can be computed by taking the weighted average of these predicted values for subjects with *X_i_* = *x*. Once all three counterfactuals are obtained, the NDE and NIE can be estimated using Equation 11 or Equation 12. We use bootstrap confidence intervals for uncertainty estimation.

The adaptability of the weighting-based approach flexibly accommodates multiple mediators works well for metabolome data. Genetic variants can affect more than one metabolite, and metabolites rarely operate independently, often jointly affecting disease risks [84]. Since weighting-based mediation analysis assumes correct specifications of models, including all metabolites (or only one or a subset of truly causal metabolites) as mediators in the outcome model may violate this assumption. This is because metabolites found to associate with a SNP may not have causal links with the disease, leading to outcome model misspecification. This issue highlights the need for a variable selection method robust to noise from potential unmeasured confounders.

Furthermore, it is naïve to assume no unmeasured confounding affects both metabolites and diseases, which is another key assumption for unbiased causal effect estimation. Sensitivity analysis using the E-value framework [85] quantifies the minimum strength of association an unmeasured confounder would need to explain away the estimated indirect effect by:

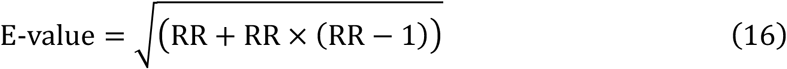

Here RR is a total natural indirect effect (TNIE) in a risk ratio. For RR *<* 1, use RR^∗^ = 1*/*RR in the above formula. A larger E-value indicates greater robustness to confounding [85].

### 4.4 Model specification using stability selection with LASSO

As the first step in estimating indirect effects for the CLSA-COM, we first fitted exposure models. For each SNP-metabolites-disease triplet, we constructed an exposure model using logistic regression. Here, the exposure *X* is the SNP, with its values indicating the number of minor alleles, and the explanatory features are the postexposure covariates *C* (age, sex, genetic principal components, hours since last meal or drink). These covariates are assumed to not be causally affected by the exposure, and therefore, treating them as covariates does not violate the sequential ignorability assumption for the identification of causal effects [86]. We then used the fitted model for computing the inverse probability weights, and the counterfactuals E[*Y_xMx_*] and E[*Y_x_*_∗_*_Mx_*_∗_] were obtained by computing the weighted averages.

Prior to fitting the outcome model, we applied stability selection with LASSO for robust selection of mediating metabolites for each SNP-metabolites-disease triplet. To validly estimate the total indirect effects using weighting-based mediation analysis, the outcome model must be correctly specified [33]. This necessitates selecting only truly relevant mediators for the outcome model. We employed stability selection with LASSO [45] for robust feature selection. In stability selection, a variable selection method, such as LASSO, is combined with bootstrapping to identify variables consistently selected across many subsamples. For each triplet, we conducted 1,000 bootstrap trials, and the metabolites with a selection probability ≥ 0.8 were included in the outcome models for causal effect estimations. The outcome model for each triplet includes a binary variable indicating a disease as the outcome (*Y*), and the mediators (*M*_1_*,…,M_k_*), covariates (*C*_1_*,…,C_l_*), SNP (*X*), and the interaction terms between the metabolites and the SNP as the independent variables. The fitted outcome model was then used for estimating the counterfactual E[*Y_x_*_∗_*_Mx_*]. LASSO analyses were performed using *R* version 4.1.3 [87] with the *glmnet* package [88].

### 4.5 Dynamic bootstrapping for computationally efficient uncertainty estimation

For the final selection of potentially causal links among SNPs, metabolites, and diseases, we conducted hypothesis tests *H*_0_ : TNIE*_t_* = 1 for all *t* = 1*,…,T* triplets using percentile bootstrap confidence intervals (PBCIs) [89] of the estimated indirect effects. To control the false discovery rate (FDR) at 5%, the PBCIs must be adjusted for multiple testing. For instance, for 483 hypotheses, the Bonferroni-adjusted significance level is *α* ≈ (0.05*/*483)∗100 = 0.0104%, requiring a (1−*α*)% confidence interval construction. The null hypothesis is rejected if this interval does not include 1, as the total natural indirect effect (TNIE) is a risk ratio. This extremely small significance level necessitates a large number of bootstrap estimates to construct a PBCI with adequate coverage. For instance, with *α* = 0.05, a 95% PBCI requires the 2.5th and 97.5th percentiles of the bootstrap distribution. With an insufficient bootstrap sample size (e.g., 10), the 2.5th percentile corresponds to the 0.25th (10*0.025) and 9.75th (10*0.975) ordered values, which do not exist. One would need at least 40 bootstrap estimates to effectively create a CI using the 1st (40*0.025) and 39th (40*0.975) ordered values, which correspond to a 100% CI, and would require an even larger sample size to construct an effective 95% CI. We estimate the causal effects separately for homozygous major vs. heterozygous (*test01*), homozygous major vs. homozygous minor (*test02*), and heterozygous vs. homozygous minor (*test12*). This results in a Bonferroni-adjusted *α* = 0.05*/*(483 ∗ 3) ≈ 0.000035. Consequently, approximately 100,000 bootstrap runs per triplet are required to construct an effective CI for all 489∗3 = 1,449 comparisons, imposing a substantial computational burden.

To reduce this computational burden, we employed a modified version of the pretesting bootstrap procedure [90] to dynamically adjust the number of bootstrap trials. We set the maximum number of bootstrap trials per triplet at *B_max_* = 120,000. This allows a maximum of two bootstrap estimates (the smallest two or the largest two) to be on the other side of 1, and the CI can still reject the null hypothesis. We first ran bootstrap estimations with an initial number of trials *B_init_* = 10,000. With 1 as the centre, if the smaller count on either side exceeds 2, the trial is terminated, and the triplet’s TNIE is considered statistically not significant. If the count does not exceed 2, we run another 1,000 bootstrap trials. This procedure continues until *B_max_* is reached. This approach effectively reduced the total number of bootstrap estimations from 120,000*483*3=174 million to 29 million (a reduction of 84%).

### 4.6 Data generation for simulation study

For our simulation study, we generated a dataset comprising the following variables: *Y*, a binary response variable representing a disease outcome. *X*, an ordinal exposure variable representing a SNP, with values indicating minor allele count. The minor allele frequency was set to 0.1. *M*_1_*,M*_2_*,M*_3_*,M*_4_*,M*_5_, continuous variables representing metabolite concentrations. We set *M*_1_ and *M*_2_ to be highly correlated (*ρ* ≥ 0.8), and we set *M*_3_*,M*_4_*,M*_5_ to be mildly correlated with each other (*ρ* ≤ 0.3). And we generated *C*_1_*,C*_2_, a binary and continuous variables (respectively), representing observed confounders such as sex and age. Finally, we generated *U*_1_, a continuous variable representing an unmeasured confounder affecting both the metabolites *M* and the outcome *Y*. The mediators were simulated by first randomly drawing *M_raw_* from the multivariate normal N(**0**,**Σ**). Here **Σ** is the covariance matrix. And we then generated each mediator by applying *M_j_* = *β_x_ X* + *β_u_U* + *M_raw_* + *ε_m_*. Here *ε_m_* ∼ N(0,1) for all *j* ∈ 1*,…,*5. This formula indicates causal relationships between *X* and the *M_j_*’s. The resulting mediator variables were then standardized to have mean zero and unit variance. The outcome variable *Y* was generated according to the model:

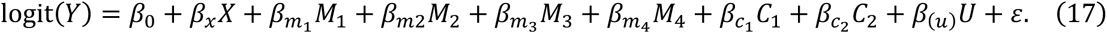

where *ε* ∼ *N*(0,1). Notably, *M*_5_ was not a true mediator, so we expected the LASSO to select only *M*_1_*,M*_2_*,M*_3_, and *M*_4_ as causal mediators. The effect sizes of the potential mediators on the outcome variable were set to 1.5, 0.8, 0.5, 0.3, respectively. We assessed the frequencies of these true causal mediators being selected under varying strengths of the unmeasured confounder (*β_u_* ∈ {0.1,0.5,0.9}), and sample sizes (*N* ∈ {100,1000,5000,8500,10000}). The simulation was run 1,000 times, and the proportion of mediators selected are calculated.

### 4.7 Quality control

Before performing mediation analyses, we conducted a series of preprocessing steps, the summary of which is illustrated in Figure 4. From the CLSA-COM, we excluded participants with a missing rate *>* 5% and those who were more closely related than second-degree relatives. Additionally, individuals whose metabolites were not measured were excluded from further analyses, resulting in a total of 8,698 subjects. Genotyping of the CLSA-COM had been conducted using the Affymetrix Axiom genotyping platform, with genetic ancestry determination performed by the CLSA group [91]. We selectively retained SNPs with a minor allele frequency (MAF) *>* 1%, a missing rate *<* 5%, and a Hardy-Weinberg equilibrium *p*-value *<* 10^−7^. With these filters, 616,294 SNPs were retained for subsequent analyses. The genotype-and individual-level QC steps were carried out using *PLINK 2.0* [79,80].

**Figure 4.**
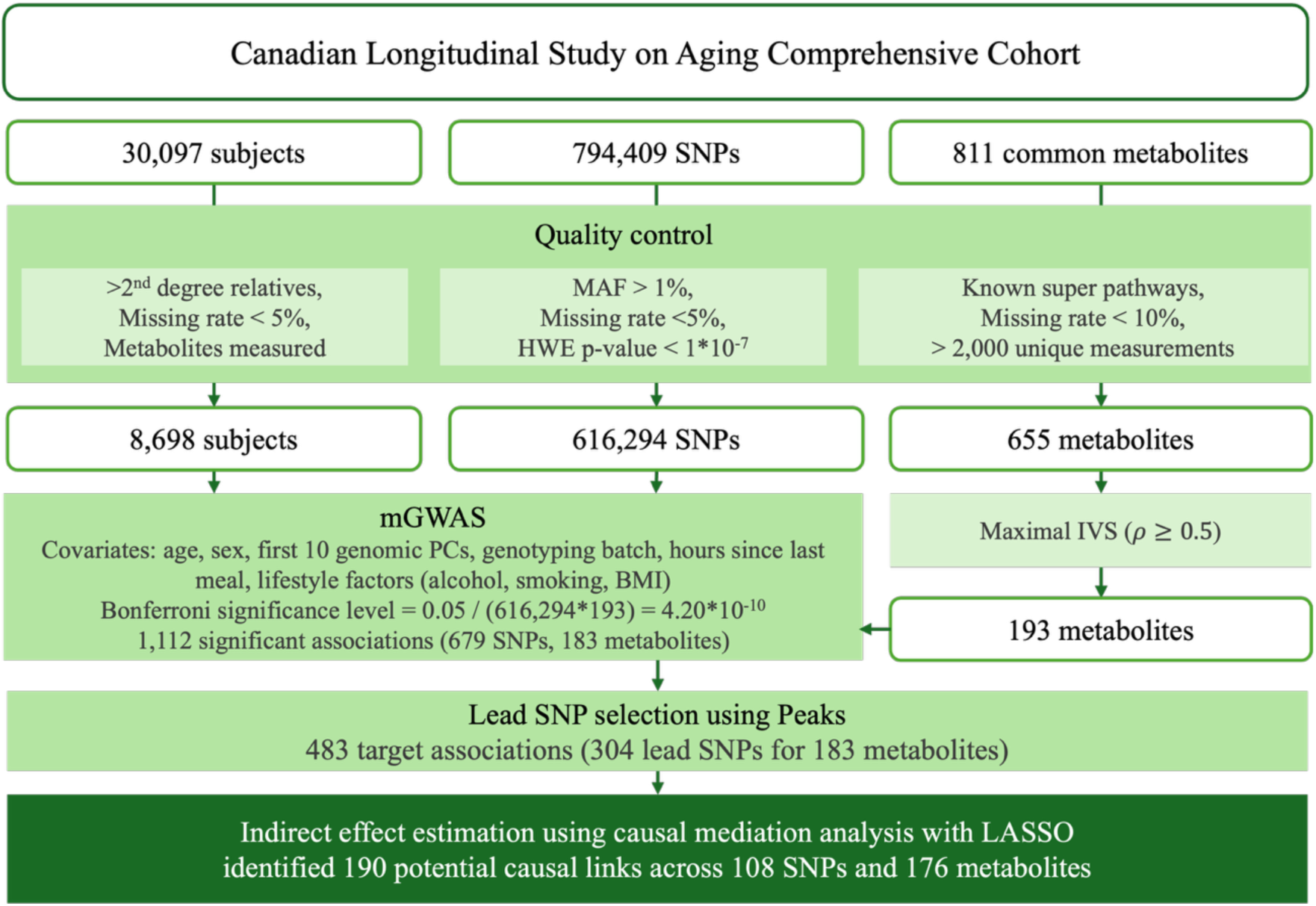
Schematic of data quality control procedures applied to the CLSA-COM dataset, and subsequent analysis steps. After a series of filtering processes, a total of 483 associations involving 304 genetic variants and 183 metabolites were selected for total indirect effect estimation using causal mediation analysis.

In the CLSA, plasma samples were quantified using ultrahigh-performance liquid chromatography-tandem mass spectrometry (UPLC-MS/MS) technology. From a total of 811 common metabolites, we included only those with known identities across eight super-pathways (amino acid, carbohydrate, cofactor and vitamins, energy, lipid, nucleotide, peptide, and xenobiotics) and with less than 10% missing values. With these filters, 655 metabolites were retained. Given the potential correlations among these metabolites, we selected maximal independent vertex sets [92] with a cutoff of 0.5, resulting in 193 independent metabolites for further analyses. Metabolome-level QC steps were carried out using *R* version 4.1.3 [87] with the *igraph* package [93].

The opinions expressed in this manuscript are the authors’ own and do not reflect the views of the Canadian Longitudinal Study on Aging.

## 5 Supplementary Information

## 5.1 Acknowledgements

This research was made possible using the data/biospecimens collected by the Canadian Longitudinal Study on Aging (CLSA; [46]). Funding for the Canadian Longitudinal Study on Aging (CLSA) is provided by the Government of Canada through the Canadian Institutes of Health Research (CIHR) under grant reference: LSA 94473 and the Canada Foundation for Innovation, as well as the following provinces, Newfoundland, Nova Scotia, Quebec, Ontario, Manitoba, Alberta, and British Columbia. This research has been conducted using the following CLSA datasets: the CLSA Baseline Comprehensive Dataset version 7.0, Genomic data version 3.0, and Metabolomics data version 2.0 under Application Number 2206033. The CLSA is led by Drs. Parminder Raina, Christina Wolfson and Susan Kirkland. The time and commitment of the participants to the CLSA study platform is gratefully acknowledged, without whom this research would not be possible.

## 5.2 Authors’ contributions

J.M. was involved in the conceptualization, data curation, visualization, and methodology of the study, also contributed with software used, conducted investigation, formal analysis, wrote, reviewed, and edited the original draft. O.V. was involved in the data curation, writing, review, and editing. A.B.-W. was involved in writing, review, editing, supervision, and project administration. L.E. was involved in writing, review, editing, supervision, and project administration.

## 5.4 Data availability

Data are available from the Canadian Longitudinal Study on Aging (www.clsa-elcv.ca) for researchers who meet the criteria for access to de-identified CLSA data.

## 5.5 Ethics approval and consent to participate

This study utilized data from the Canadian Longitudinal Study on Aging (CLSA). The CLSA obtained written and informed consent from all participants during the original data collection [46]. All data used for this analysis were de-identified and accessed through the CLSA data access application process. The specific analytical protocol for this study was reviewed and approved by the Joint Clinical Research Ethics Board of BC Cancer, the University of British Columbia, and Simon Fraser University. The assigned Research Ethics Board (REB) certificate number is H22-01012.

